# Is there a higher risk of exposure to *Coxiella burnetii* for pre-clinical veterinary students?

**DOI:** 10.1101/2022.04.22.22274162

**Authors:** Anne Conan, Christa Gallagher, Nicole Erskine, Michael Howland, Marshalette Smith-Anthony, Silvia Marchi, Ioannis Magouras, Ananda Muller, Anne AMJ Becker

## Abstract

*Coxiella burnetii* is globally distributed but evidence of zoonotic transmission in the Caribbean region is scarce. The presence of the bacterium is suspected on the Caribbean island of St. Kitts. The risk of exposure of veterinary students was reported in other regions of the world but is not documented in the Caribbean region. The present study aimed to evaluate the risk of exposure to *C. burnetii* for pre-clinical veterinary students attending an island-based veterinary school. A cross-sectional study was conducted to compare incoming and outgoing veterinary students’ seroprevalence. Serology was performed using indirect immunofluorescence assay to test *Coxiella burnetii* Phase I and Phase II immunoglobulins M and G. Background data were gathered using a standardized questionnaire.

Of the 98 participants (48 incomings and 50 outgoings), 41 (41.8 %, 95 %CI: 31.9-52.2) were seropositive to *C. burnetii*. There was no significant difference between the two groups (45.8% for incoming vs. 38.0% for outgoing) (p=0.4). No risk factors were significantly more reported in the seropositive group. U.S. state of origin of the students was not associated with seropositivity either.

Pre-clinical veterinary students do not have a higher risk of exposure to *C. burnetii* by attending the veterinary school in St. Kitts, but they are highly exposed before arrival on the island. Most of these participants had experience with animals either through farming or previous veterinary technician employment. This indicates a high exposure in the U.S. young population aiming to become veterinarians. There is an urgent need to increase *C. burnetii* surveillance in animals and humans to apply relevant prevention and control measures, including recommendations for vaccination of students and professionals at risk.

**Author Summary:** Q fever is a zoonosis present worldwide. Risk of exposure is higher in livestock farmers and other animal-related professions, including veterinary students. The presence of the bacterium is suspected on the Caribbean island of St. Kitts. We aimed to evaluate the risk of exposure to *C. burnetii* for students attending an American veterinary school based in St. Kitts. By comparing seroprevalence in incoming and outgoing students, we observed no risk of exposure by attending the school. These findings are different from studies in other veterinary schools. Future studies could explain these geographical differences and inform veterinary schools worldwide on appropriate measures to protect students from exposure to *C. burnetii*. Moreover, we detected a high seroprevalence in the incoming population, proving an anterior exposure. This indicates a high exposure in the U.S. young population aiming to become veterinarians. There is an urgent need to increase *C. burnetii* surveillance in animals and humans to apply relevant prevention and control measures, including recommendations of vaccination of students and professionals at risk.

## Introduction

*Coxiella burnetii* is an obligate intracellular bacterium responsible for the zoonosis Q fever [1]. All mammalian species including humans and some bird species can be infected by the bacteria. The infection of animals is usually asymptomatic or manifests as mild fever, with sporadic abortions in late pregnancies or moribund offspring [2]. In humans, most infections are also asymptomatic. Acute Q fever is characterized by an influenza-like illness. However, a more severe form with pneumonia or hepatitis may occur. Some individuals may also develop a chronic infection with risk of endocarditis [1]. During infection, the bacterium undergoes a phase of transition, leading to the presence of two serologically distinguishable phases. Phase I type is the bacterium’s virulent form, and Phase II is avirulent. The diagnosis of Q fever in humans is based on the association between clinical signs and serological status. The course of the disease is serologically divided into three stages: the onset with high Phase II immunoglobulins IgG and IgM, the acute phase with higher Phase II IgG, and a chronic phase characterized by high Phase I IgG [3].

*Coxiella burnetii* is considered endemic worldwide with the exemption of New Zealand. Infections are usually sporadic, but major outbreaks in animals and humans have occurred in Australia [4] and The Netherlands [5]. Outside of these outbreaks, human Q fever is regarded an occupational disease, with a higher risk of exposure in livestock farmers, abattoir workers, and veterinarians. Veterinary students are also at risk, with reported seroprevalence of 22 % in Brazil [6], 35 % in Iran [7], 20 to 30 % in The Netherlands [8], up to 17 % in Spain [9] and 17 to 58 % in Slovakia [10].

In the U.S., the prevalence of *C. burnetii* in livestock and the risk of human exposure are considered low. Two old studies (1964 and 1974) reported a seroprevalence of 5 and 10 % in veterinary students, respectively [6,11]. More recently however, a national investigation reported a high exposure in veterinarians (22 %) [12], highlighting the need for further investigations in the veterinary curriculum and profession.

Ross University School of Veterinary Medicine (RUSVM) delivers a pre-clinical Doctor of Veterinary Medicine (DVM) curriculum in St. Kitts (West Indies). Evidence of *C. burnetii* in the Caribbean is limited, with few reports from several islands, including Trinidad, Grenada, and Cuba [13–15]. In St. Kitts, several studies reported seropositive humans or animals [16–18]. The main objective of the study was to evaluate the risk of exposure to *C. burnetii* for students attending RUSVM in St. Kitts by comparing the seroprevalence of on-island incoming students and outgoing students.

## Material and Methods

### Target population

At RUSVM, undergraduate students enter the school either in 1^st^ semester or in a 15-week Veterinary Preparatory program. The curriculum is accelerated and one semester lasts four months (Spring, Summer and Fall semesters). After seven ‘semesters’ (∼two and half years) of pre-clinical coursework, the students complete their program with a three-semester clinical curriculum at an affiliated school, usually based in the U.S.

### Enrolment of participants

Inclusion criteria for the participants was the registration at RUSVM in the Veterinary Preparatory program or first semester for incoming group and in seventh semester for outgoing group. In January 2020, the targeted population varied between 140 (outgoing) to 183 individuals (incoming). Fifty individuals were randomly selected in both groups based on the registration list of the Registrar’s Office. The randomisation was performed in Microsoft Excel™ using the function ‘RANDOM’. If an invitee declined the participation or did not confirm participation after six days, the next individual on the random list was invited to participate. Exclusion criteria for incoming students were their presence in St. Kitts for more than a month (based on air plane arrival) or a previous visit/vacation in St. Kitts.

### Enrolment changes due to COVID-19 pandemic

Enrolment started in January 2020. Unfortunately, the teaching program at RUSVM was disrupted from mid-March 2020 onwards due to the COVID-19 pandemic and consequent travel restrictions. Online teaching was initiated and most of the students left St. Kitts to continue the curriculum from home. Face-to-face activities on island gradually resumed from September 2020, starting with 7^th^ semester, until September 2021 (Veterinary Preparatory). To decrease temporal bias, incoming student enrolment was adapted from September 2020. Inclusion criteria were extended to registration in second or third semester at RUSVM. Exclusion criteria (less than a month in St. Kitts and no previous visit of St. Kitts) remained the same.

### RUSVM employees

In parallel with the student investigation, we provided the opportunity for RUSVM employees that were in contact with livestock on campus to get tested for antibodies. Contact with livestock was defined as work with the RUSVM farming animals (cattle and sheep) at less than 5 meters for more than 30 minutes at least once a month or with opened carcasses of farming animals at RUSVM during the last 6 months. Due to COVID-19 pandemic, exclusion criterium of being abroad for more than a month in the last six months was added. Department heads provided the list of employees who fit the above criteria, and those employees were sent invitations to participate in the study. Forty-one employees fit our definition and were invited to join.

### Questionnaire

Students responding positively to the study invitation email were scheduled an appointment. After signing a consent form, a standardized questionnaire was verbally administered to the participants and directly entered with Qualtrix®. Questions included the possible associated factors to *C. burnetii* seropositivity as follows: demographic factors, living area, animal ownership, past veterinary/animal work, consumption of raw milk, and exposure to ticks. The outgoing students were also asked about their extra-curricular activities with animals during their time in St. Kitts. The questionnaire is available in Appendix 1.

### Sampling

A blood sample of maximum 8 mL was drawn by venipuncture by a certified nurse at Health Services of RUSVM. All blood collection tubes were centrifuged within 30 minutes to 3 hours after collection and the serum was stored at -20°C.

### Serological testing

Two commercially available indirect immunofluorescent assays (IFA) were used to screen all samples for human IgG and IgM antibodies to *C. burnetii* (Focus Diagnostics Q Fever IFA IgG and IgM assays, Cypress, CA). Both immunoglobulin-specific assays consisted of slide wells in which each well contained 2 individual spots with *C. burnetii* (Nine Mile strain) Phase I and Phase II antigens, respectively. The human positive and negative control samples (Focus Diagnostics Q Fever ref IF0211 and IF0213) served as the reference markers in identifying positive and negative results. The presence of bright green fluorescence of coccobacillary morphology and lack of background fluorescence were used to identify positive samples, while the total absence of fluorescence identified negative samples. All serum samples were initially screened at the manufacturer’s recommended serum dilution (1:16) with the provided IgG Sample Diluent (Figure 1A and 1B). Any serum sample found to be positive at the screening dilution for both IgG Phase I and II was further titrated in reconstituted phosphate buffered saline (PBS) using the manufacturer’s recommended serum dilutions of 1:64, 1:128, 1:256, 1:512 and 1:1024 (Figure 1C). Any serum sample found to be positive for only one Phase of IgG was taken to end titer by threefold serial dilutions. Likewise, serum samples were screened for IgM antibodies at the manufacturer’s recommended serum dilution (1:16) with IgM pre-treatment diluent and subsequently diluted three- or six-fold to determine end titers (Figure 1D). All IFAs were consistently performed by the same person, as well was the interpretation of all slides by another person. Both were blinded for the sample group (incoming, outgoing, employees). A random number of slide images were additionally sent to the German National Consiliary Laboratory of *Coxiella burnetii* (Stuttgart, Germany) for confirmation.

**Figure 1:**
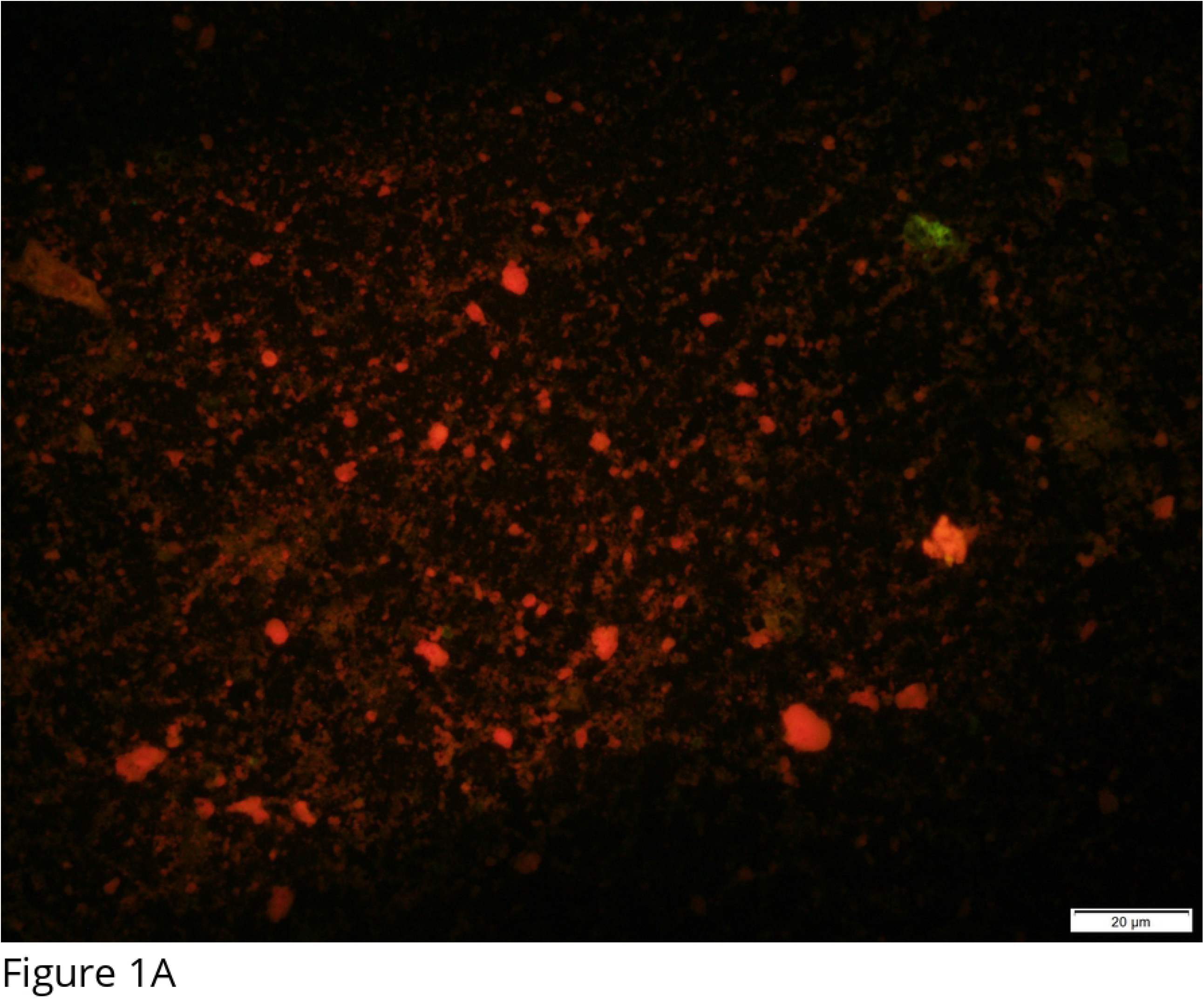
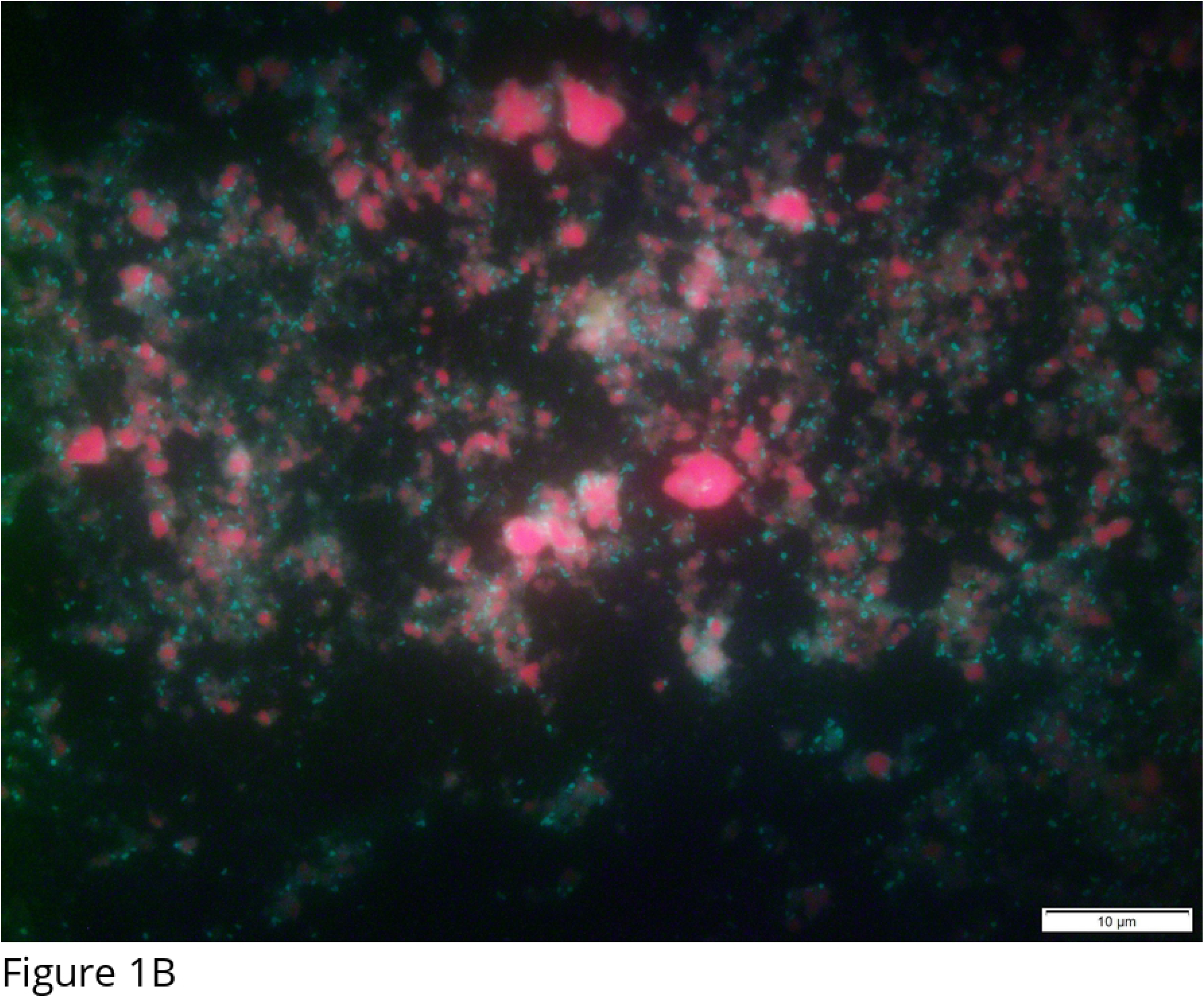
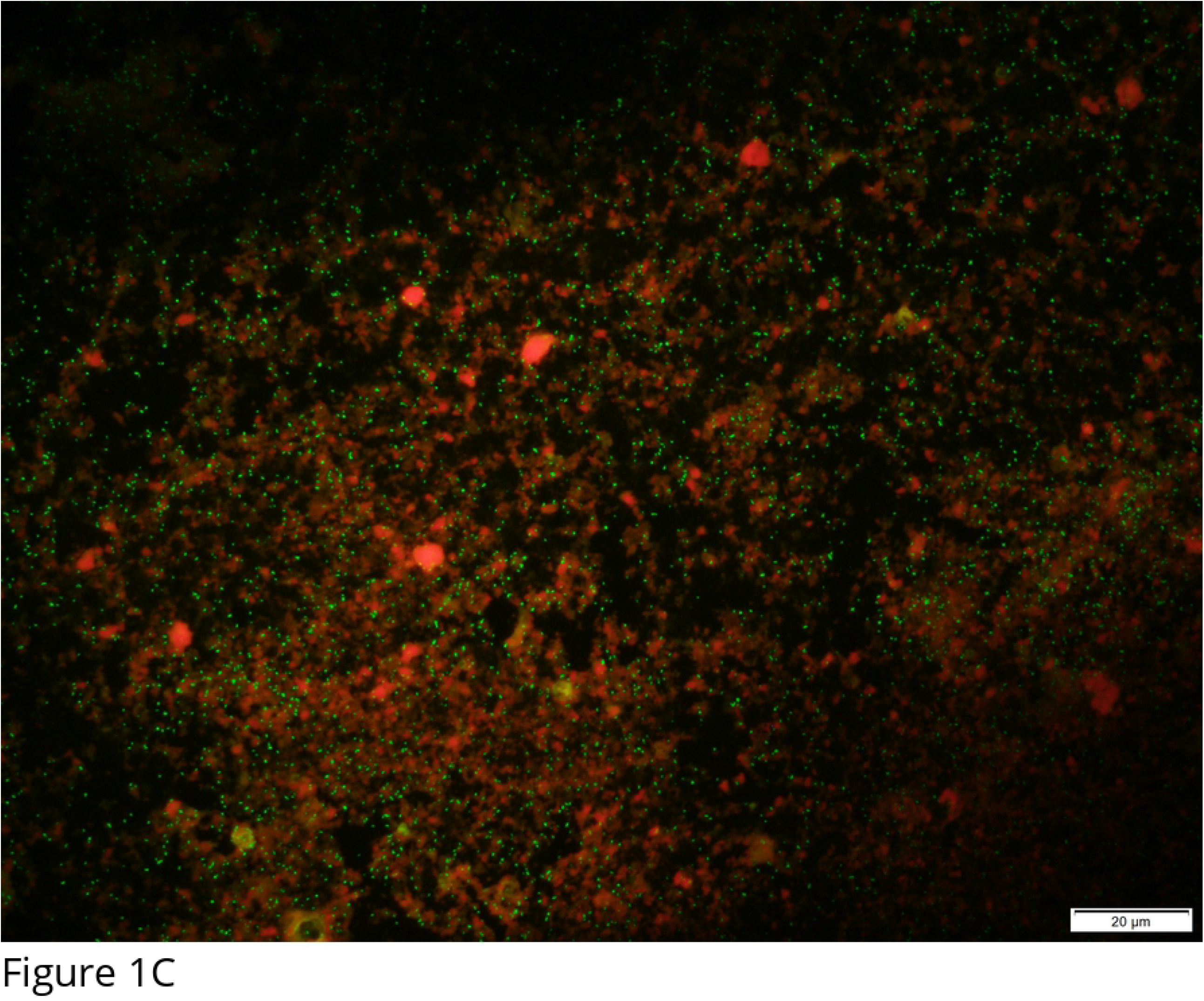
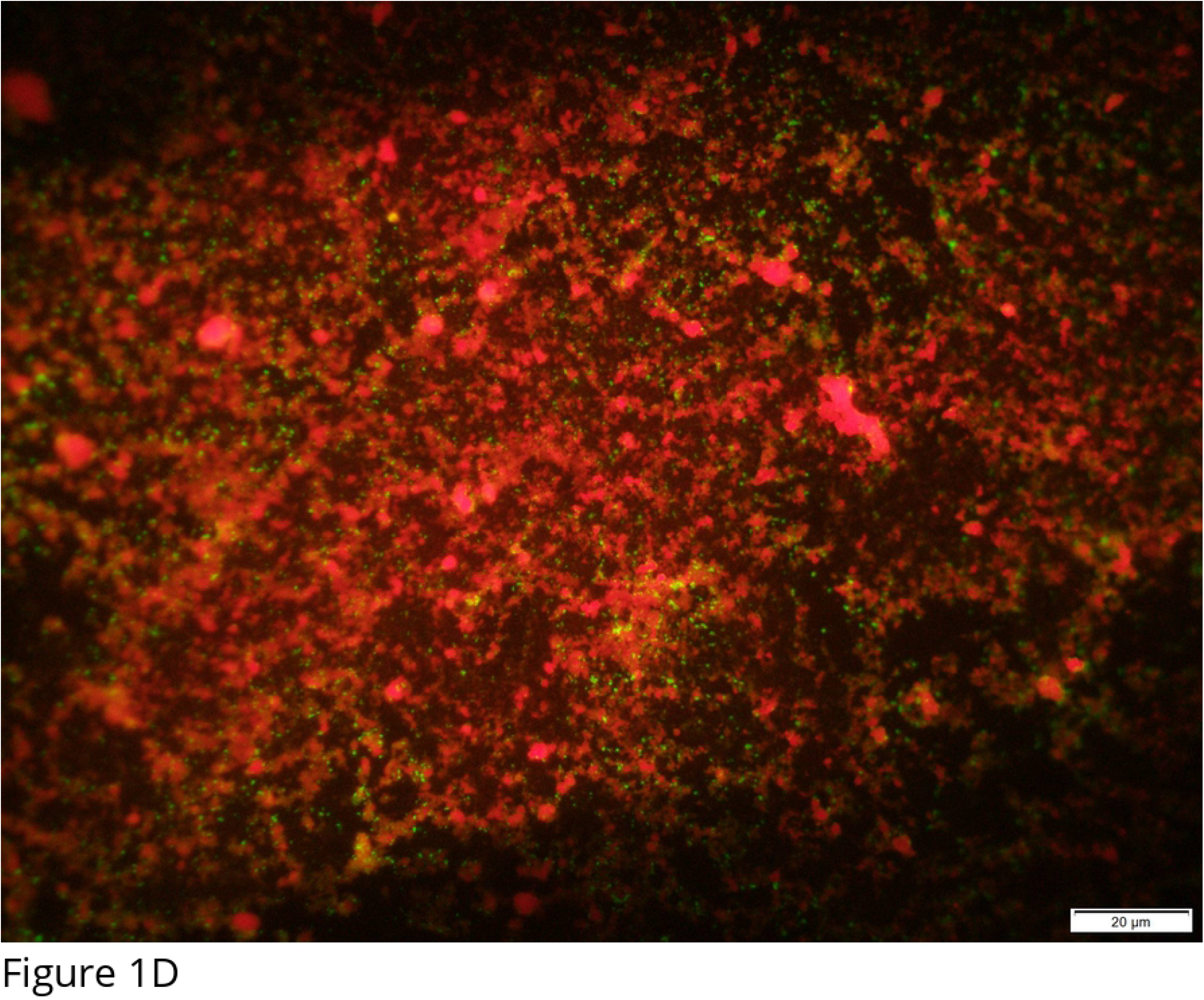
Examples of results for the indirect immunofluorescent assay A) Negative; B) Positive for IgG screening at dilution 1:16; C) Positive for IgG dilution 1:64; D) Positive for IgM dilution 1:64

### Case definition

Our study did not record symptoms as the objective was to investigate the exposure. An individual with any immunoglobulin concentration equal or above the threshold of 1:64 was classified as seropositive. Cases of acute and chronic exposures were also defined and reported to participants. Acute exposure was suspected when Phase I and/or Phase II IgG were equal or superior to 1:256 and 1:512, respectively. A high phase I IgG equal or above 1:512 and superior to Phase II IgG would lead to suspicion of chronic infection. Finally, individuals with negative IgG and positive Phase II IgM (superior to 1:64) were suspected of developing an early infection [19–21].

### Data analyses

Seroprevalence was expected to be 5 % for incoming students and 20 % for outgoing students, respectively. To detect a difference with a power of 80 % and a significance of 5 %, 50 individuals needed to be sampled in each group. All data were merged in a unique database and analysed using R. Confidence intervals of 95 % (95% CI) were computed based on the binomial distribution. The main exposure variable (incoming/outgoing) was tested by logistic regression. Other variables were compared using logistic regression with the incoming/outgoing variable forced in the model. For the sub-population of outgoing students, activities performed during their stay in St. Kitts were also tested.

Among the tested factors, origin location was used in different ways. We collected information about all world locations where the participant lived for over a year. Residence location scale was the state for the U.S. and province for Canada, and the country for the rest of the world. As most participants previously lived in the U.S., only this location information was used for risk factor analysis. We grouped the states by population of cattle, sheep and/or goats (based on 2019 census, USDA, https://quickstats.nass.usda.gov/) and by the number of human Q fever cases reported in 2019 (CDC data, https://www.cdc.gov/qfever/stats/index.html). Each created variable was binary using the median as the threshold between high and low risk. Absence of data in a state was considered as no livestock population. An individual who lived in two states differing in risk (e.g. one state with no cattle and one state with high number of cattle) was attributed with the highest risk value (variable=1). All created variables were tested using a logistic regression with the incoming/outgoing variable forced in the model.

### Ethic statement

The ethic approvals were granted by (i) Interim Ethic Review Committee (IERC, Ministry of health, community development, gender affairs and social services, St. Kitts; protocol #: IERC-2020-01-037), (ii) the institutional review board of RUSVM) (IRB protocol #19-05-XP) and (iii) the Human Subjects Ethics Sub-Committee of City University of Hong Kong (protocol #: 41005-20 Conan, application no H002441A)

## Results

Out of the 98 enrolled participants (incoming and outgoing), 41 (41.8 %, 95 % CI: 31.9-52.2) were considered as seropositive to *C. burnetii*. The seropositivity of outgoing students was not significantly different from the seropositivity of incoming students (p=0.4) (Table 1). Seroprevalence in employees was high with 8 seropositives out of 15 sampled individuals (53.3 %, 95 %CI: 26.6-78.7).

**Table 1:**
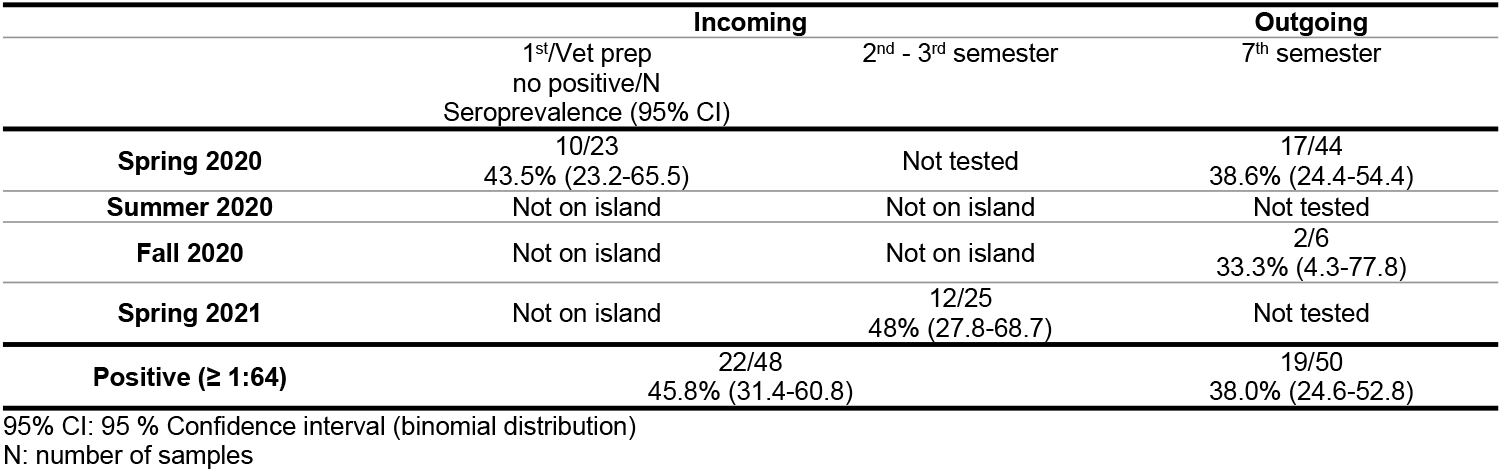
Seroprevalence and 95 % confidence interval of the incoming and outgoing students in St Kitts(case definition: at least one immunoglobulin dilution equal or above 1:64). Seroprevalence by enrolment group is presented

Distribution of dilution by phase is presented in Table 2. Dilution was not performed until negativity, so only thresholds at 1:128 (Phase I) and 1:64 (Phase II) are described. There were more individuals positive to Ig Phase II (n=38) than to Ig Phase I (n=26, McNemar’s test p=0.002). There were no other differences between particular immunoglobulins (Table 2). None of the tested factors were significant during univariate analysis (p>0.05) (Table 3 and Appendix 2). Looking at the outgoing participants only, none of the tested exposure factors linked with their activities at RUSVM were different between seropositive and seronegative individuals. No significant association between student club or live animal activities and seropositivity in outgoing participants were found (Appendix 3).

**Table 2:**
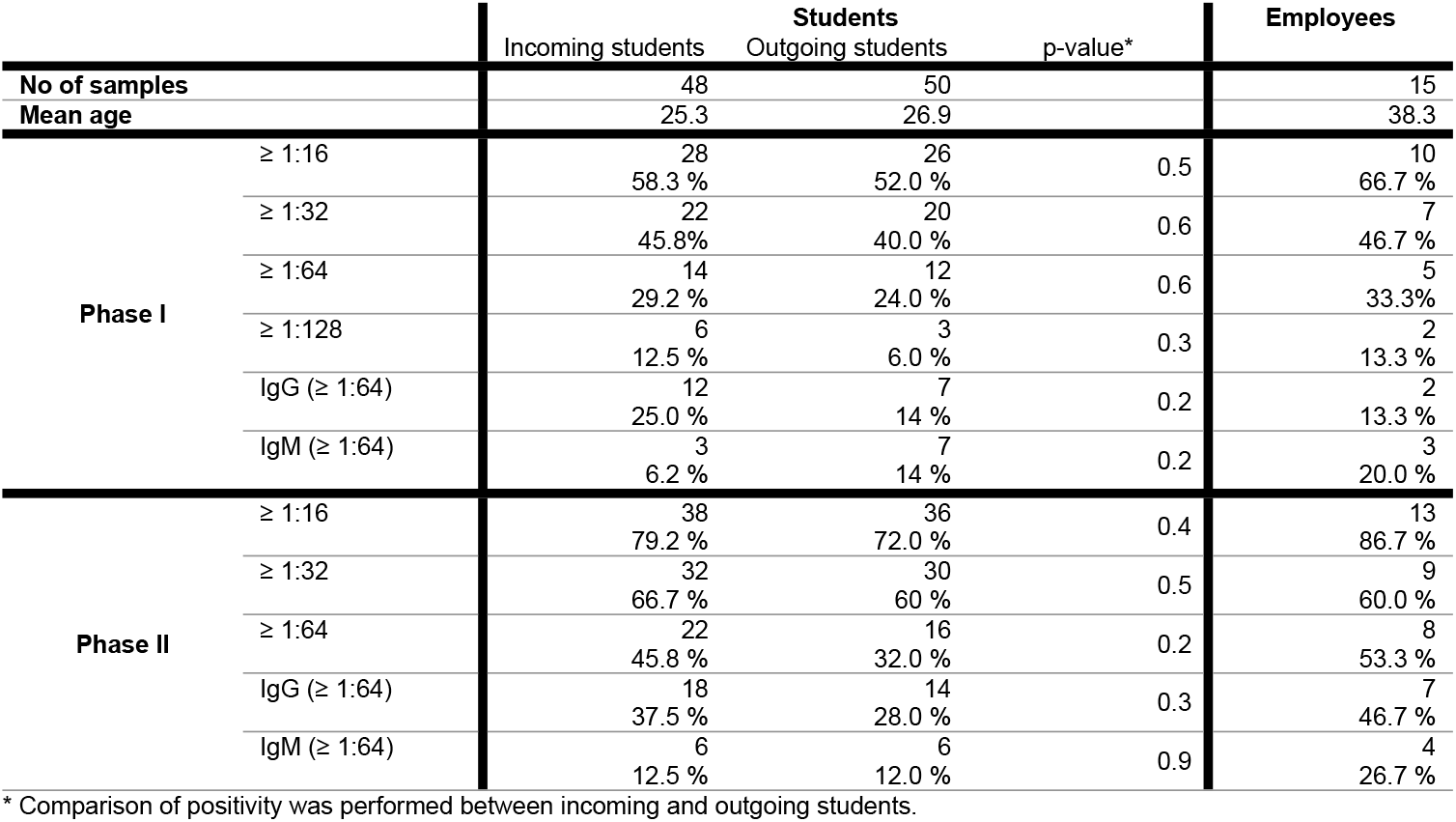
Number and proportions of samples by phase and by positive dilution in the three groups (incoming, outgoing and employees)

**Table 3:**
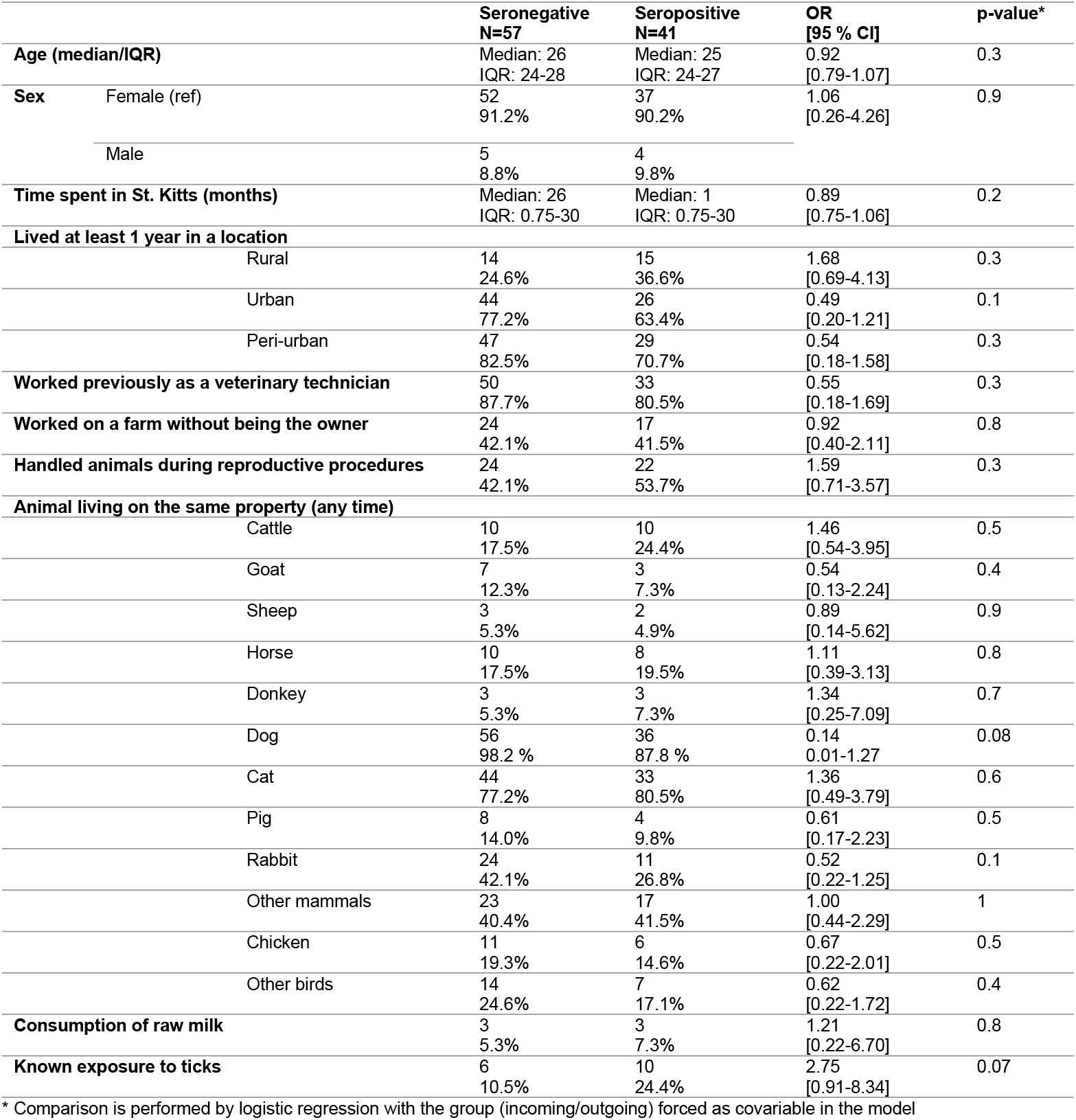
Exposure factors for Coxiella burnetii seropositivity in the student population.

|  | Seronegative<br>N=57 | Seropositive<br>N=41 | OR<br>[95 % CI] | p-value* |
| --- | --- | --- | --- | --- |
| **Age (median/IQR)** | Median: 26<br>IQR: 24-28 | Median: 25<br>IQR: 24-27 | 0.92<br>[0.79-1.07] | 0.3 |
| **Sex** |  |  |  |  |
| Female (ref) | 52<br>91.2% | 37<br>90.2% | 1.06<br>[0.26-4.26] | 0.9 |
| Male | 5<br>8.8% | 4<br>9.8% |  |  |
| **Time spent in St. Kitts (months)** | Median: 26<br>IQR: 0.75-30 | Median: 1<br>IQR: 0.75-30 | 0.89<br>[0.75-1.06] | 0.2 |
| **Lived at least 1 year in a location** |  |  |  |  |
| Rural | 14<br>24.6% | 15<br>36.6% | 1.68<br>[0.69-4.13] | 0.3 |
| Urban | 44<br>77.2% | 26<br>63.4% | 0.49<br>[0.20-1.21] | 0.1 |
| Peri-urban | 47<br>82.5% | 29<br>70.7% | 0.54<br>[0.18-1.58] | 0.3 |
| **Worked previously as a veterinary technician** | 50<br>87.7% | 33<br>80.5% | 0.55<br>[0.18-1.69] | 0.3 |
| **Worked on a farm without being the owner** | 24<br>42.1% | 17<br>41.5% | 0.92<br>[0.40-2.11] | 0.8 |
| **Handled animals during reproductive procedures** | 24<br>42.1% | 22<br>53.7% | 1.59<br>[0.71-3.57] | 0.3 |
| **Animal living on the same property (any time)** |  |  |  |  |
| Cattle | 10<br>17.5% | 10<br>24.4% | 1.46<br>[0.54-3.95] | 0.5 |
| Goat | 7<br>12.3% | 3<br>7.3% | 0.54<br>[0.13-2.24] | 0.4 |
| Sheep | 3<br>5.3% | 2<br>4.9% | 0.89<br>[0.14-5.62] | 0.9 |
| Horse | 10<br>17.5% | 8<br>19.5% | 1.11<br>[0.39-3.13] | 0.8 |
| Donkey | 3<br>5.3% | 3<br>7.3% | 1.34<br>[0.25-7.09] | 0.7 |
| Dog | 56<br>98.2 % | 36<br>87.8 % | 0.14<br>0.01-1.27 | 0.08 |
| Cat | 44<br>77.2% | 33<br>80.5% | 1.36<br>[0.49-3.79] | 0.6 |
| Pig | 8<br>14.0% | 4<br>9.8% | 0.61<br>[0.17-2.23] | 0.5 |
| Rabbit | 24<br>42.1% | 11<br>26.8% | 0.52<br>[0.22-1.25] | 0.1 |
| Other mammals | 23<br>40.4% | 17<br>41.5% | 1.00<br>[0.44-2.29] | 1 |
| Chicken | 11<br>19.3% | 6<br>14.6% | 0.67<br>[0.22-2.01] | 0.5 |
| Other birds | 14<br>24.6% | 7<br>17.1% | 0.62<br>[0.22-1.72] | 0.4 |
| **Consumption of raw milk** | 3<br>5.3% | 3<br>7.3% | 1.21<br>[0.22-6.70] | 0.8 |
| **Known exposure to ticks** | 6<br>10.5% | 10<br>24.4% | 2.75<br>[0.91-8.34] | 0.07 |

\* Comparison is performed by logistic regression with the group (incoming/outgoing) forced as covariable in the model

Interestingly, the four participants suspected of acute infection (2 incoming, 1 outgoing and 1 employee) had different backgrounds. Three were females and one was male. While the employee lived exclusively in St. Kitts, the students lived in different states in the U.S. (Michigan, Minnesota, North Dakota, Ohio, Pennsylvania, Utah) and in different environments (rural N=3, peri-urban N=0, urban N=3). Two had previous experience as veterinary technicians with ruminants and two had worked on farms. In St. Kitts, the incoming participants lived on campus and two others in two different island parishes. Species of animals owned by these four participants also varied among them (cattle N=2, sheep N=2, donkey N=1, cat N=3, pig N=2, rabbit N=1). All have owned a dog but three of them only in the last year. None of the four have ever consumed raw milk and one reported to have been bitten by a tick. The participants showing serological evidence of early infections were four incoming, two outgoing and one employee, all females. However, six out of seven had experience as veterinary technicians with pets and four indicated previous work on a cattle farm. None of the participants fit our definition of chronic infection.

## Discussion

The seroprevalence in the population of students outgoing RUSVM was not significantly different from the seroprevalence of the incoming students. Therefore, studying pre-clinical veterinary sciences in St. Kitts doesn’t increase the risk of being exposed to *C. burnetii*. These results differ from what was observed in Spain [9] or in The Netherlands [8,22]. In these veterinary schools, the risk of being seropositive to *C. burnetii* increased with the number of years spent at the university. Several hypotheses could explain the difference in our results. First, RUSVM has a pre-clinical curriculum and therefore, students have less contact with live animals compared to their subsequent clinical years. However, RUSVM students are exposed to live animals through different laboratory sessions that include manipulation of sheep and cattle, and also through extracurricular activities. Secondly, the prevalence of *C. burnetii* in livestock could be lower in St. Kitts compared to European countries. While the seroprevalence in a male sheep flock was found to be 26.3 %, the prevalence and prevalence of animal shedding are unknown [18]. Finally, the exposure environment may differ with animals free-roaming in the community and general animal manipulations occuring outdoors, therefore preventing concentration of the bacteria in the environment. This, associated with the application of basic personal protective equipment may lower the transmission risk to students. Different studies, such as follow-up serology on RUSVM students during their clinical year, a prevalence estimation in St. Kitts’ livestock and testing closed environments in veterinary schools could support these hypotheses. Such results could inform veterinary schools worldwide on appropriate measures to protect students from exposure to *C. burnetii*.

Although the seroprevalence is not different between the two student groups, the baseline in incoming students (43.5%) is higher than we expected. Two old studies (1960s-70s) detected seroprevalence of 5 % in veterinary schools in the U.S. Our results indicate that there is a high risk of exposure in this young population. We cannot generalize to the entire U.S. population, as veterinary students tend to have a history of more frequent and diverse contact with animals. Most study participants had indeed contact with animals through internships, veterinary technician work or farming prior to enrolment in the veterinary school. Still, the seroprevalence observed is higher than previously reported in the veterinary profession (22.2%) [12]. Seroprevalence among healthy adults has been estimated in the U.S. at 3.1 % with higher rates in males than females [23]. Our data are closer to the 49 % seroprevalence reported in veterinarians in Nova Scotia (Canada) [24]. Also, our results don’t show a specific location pattern: no states or regions (based on geography, livestock population, or Q fever human cases) show a significant association to seropositivity. Finally, it is almost certain that the infection is underreported in animal and human populations [18,25]. Several authors have already mentioned a possible increase of prevalence in the U.S.[25,26]. Therefore, our results should urge human and veterinary authorities to improve U.S national surveillance and increase awareness of animal-related workers. These measures would help with early detection, risk communication, risk management, and prevention of severe outbreaks similar to those reported in The Netherlands or in Australia.

In our risk factor analysis of *C. burnetii*, none of the tested factors showed a significant difference between seronegative and seropositive participants. While the power of the exposure analysis may be too low for this study (as the sample size was calculated for a difference of seroprevalence between groups) and there is a certain recall bias, some trends can be observed and hypothesis posed. First, most of the participants (seronegative or seropositive) worked previously as veterinary technicians which confirms the population particularity of our target population compared to the general population. Second, four factors were more frequent (but not significant) in seropositives: known exposure to ticks, residence location in rural areas, reproductive procedures (particularly in cattle) and working as a cattle veterinary technician.

These factors are known to be risk factors of *C burnetti* exposure. Usually, risk factors on the American continent would include living on a farm [12,27]. In the Netherlands, the veterinary student seropositivity was associated with the number of years lived on farms [8]. Interestingly, the factors “owning a dog” and “work on an equine farm” were more frequent in seronegatives. Further investigation focusing on these risk factors should be performed to better understand the risk of transmission of *C. burnetii*. Finally, no geographical difference could be observed. This would indicate that the infection is actually endemic in most of the U.S. territory. It is possible that the state differences in CDC-reported human case result from better surveillance in some states compared to others.

The employee population was not added to our exposure factor analyses because of the high disparity in their demographics, country origin, and professional background compared to a homogenous student population. We also observed high seroprevalence in this population (53.3 %) but due to the low sample size no conclusions can be made about potential risk factors. However, this confirms the risk of exposure to *C. burnetii* in the veterinary and related professions [1]. A larger seroprevalence study could be conducted in veterinary school workers in North America and other regions to identify potential risk activities. Awareness of animal workers should be raised and regular testing recommended. Moreover, the provision of vaccination should be considered for staff working in veterinary and agricultural schools where the employees are in regular contact with animals.

In total, four participants were suspected to be in the acute form of the disease, but none of the participants fits the definition of chronic disease. These four participants were encouraged to visit the RUSVM Health Services or their personal doctors. One of the study limitations is the absence of questions related to current or recent symptoms. The detection of acute disease followed by appropriate treatment could reduce the risk of progression to chronic disease. Therefore, awareness campaigns for veterinary students, veterinary-related workers, and human practitioners to increase the testing of *C. burnetii* in case of flu-like symptoms is recommended. Other limitations of our study include the lag between incoming participant arrival and sampling and the IFA interpretation. First, the incoming students tested in the month after arrival could have been exposed at arrival on St. Kitts. Indeed, IgM can be detected within two weeks after exposure [28,29]. However, this hypothesis is unlikely as the comparisons between group by phase and immunoglobulin were not significant. The IgG seroprevalence would be low if there was exposure during this first month [30]. Also, the incoming students have no direct animal contact during the first month of the curriculum. Moreover, the 2^nd^ cohort of incoming participants arrived in St. Kitts while COVID-19 restrictions were still in place and were therefore quarantined in their accommodation for two weeks.

The second limitation is the IFA test, that can be prone to subjective interpretation. We implemented several safeguards to decrease this bias: the high cut-off of 1:64 (also avoiding cross-reactions with Rickettsia), consistent interpretation by a singular, and a random confirmatory reading by the German National Consiliary Laboratory of *Coxiella burnetii*. Therefore, we believe the bias of IFA interpretation has been minimized.

In conclusion, our study does not indicate an increased risk of exposure to *C. burnetii* for students attending RUSVM. However, we report a high seroprevalence in the incoming student population, indicating a high risk of exposure in the U.S. young population aiming to become veterinarians. This highlights the need for urgent and necessary measures to increase surveillance of *C. burnetii* in the human and animal populations in the U.S. Measures of prevention and control, and awareness of the animal-worker population should be improved, and vaccination of animals and humans should be considered at the farm, veterinary school, and national levels.

## Data Availability

The data that support the findings of this study are available on request from the corresponding author (AC). This includes only deidentified (names, date of enrolment, semester and origin data removed) student data. Data from RUSVM employees will not be shared. The data are not publicly available due to ethical restrictions. The containing information could compromise the privacy of research participants.

## Acknowledgement

We are grateful to Mary Cartwright (Research Assistant), Juliet Battice (Student Health Services) and Shianne England (Student Health Services) for helping with the data collection. We thank Dr. Larissa Dangel (German National Consiliary Laboratory of Coxiella burnetii, Stuttgart, Germany; State Health Office Baden-Württemberg, Stuttgart, Germany) for her support. AC, CG, MSA, AB obtained financial support by the One Health Center for Zoonoses & Tropical Veterinary Medicine, Ross University School of Veterinary Medicine, St. Kitts and Nevis (Intramural grant # 41005-20). The funding source had no involvement in the study design, in the collection, analysis, and interpretation of data, in the writing of the paper or the decision to submit the paper for publication.

## Authors’ contributions

Conceptualisation: AC; Methodology: AC, MSA, AB; Supervision: AC, CG, AM; Formal analysis: AC, AB, Funding acquisition: AC, CG, MSA, AB; Investigation: NE, MH, MSA, SM; Writing – Original draft: AC; Writing - review &editing: CG, IM, AM, AB

---

Appendix 1: Standardized questionnaire administered to all study participants.

Appendix 2: Exposure factors for *Coxiella burnetii* seropositivity in the student population

(Continued Table 3)

Appendix 3: Exposure factors for *Coxiella burnetii* seropositivity in the outgoing student population

